# Identification of Suicide-Related Subgroups Using Latent Class Analysis: Complementary Insights to Explainable AI–Based Classification

**DOI:** 10.64898/2026.03.25.26349264

**Authors:** Busenur Kizilaslan, Lars Mehlum

## Abstract

**Purpose:** Suicide and self-harm are major public health concerns characterized by substantial clinical and psychosocial heterogeneity. While latent class analysis has been used to identify subgroups of people with suicidal behavior, the extent to which such population-level phenotyping complements explainable artificial intelligence–based classification models remain unclear.

**Methods:** We applied latent class analysis to a cross-sectional, publicly available dataset of 1000 individuals presenting with self-harm and suicide-related behaviors at Colombo South Teaching Hospital, Kalubowila, Sri Lanka. Sociodemographic, psychosocial, and clinical variables were used to identify latent subgroups. Class characteristics and suicide prevalence were examined and compared with variable importance patterns reported in a previously published explainable artificial intelligence (XAI)-based suicide classification study using the same dataset.

**Results:** Four latent classes were identified. Two classes exhibited very high suicide prevalence (91.2% [95% CI: 87.7– 93.8] and 99.0% [95% CI: 96.4–99.7]), whereas two classes showed low prevalence (<1%). The two high-prevalence classes differed markedly in lifetime psychiatric hospitalization history, with one class showing a 100% prevalence of prior hospitalization and the other substantially lower hospitalization rates. These patterns partially aligned with, and extended beyond, variable importance findings from the XAI-based model.

**Conclusion:** Latent class analysis identified distinct subgroups with substantially different suicide prevalence and clinical profiles, underscoring the heterogeneity of individuals presenting with self-harm. Comparison with XAI-based suicide classification model findings suggest that unsupervised phenotyping and supervised classification provide complementary perspectives, offering population-level context that may enhance the interpretability of suicide assessment frameworks.

## Introduction

Suicide and self-harm are complex and multifactorial phenomena shaped by demographic, psychosocial, and clinical factors, resulting in substantial heterogeneity in risk profiles that complicate assessment and intervention. In recent years, machine learning (ML) and explainable artificial intelligence (XAI)–based models have shown promising performance in estimating individual suicide risk [1, 2]. However, these supervised approaches primarily generate individual-level predictions and may not fully capture population-level patterns or latent subgroup heterogeneity.

Latent class analysis (LCA) is a probabilistic, model-based method that identifies unobserved subgroups of individuals based on shared patterns of variable characteristics while explicitly accounting for uncertainty in class membership. In suicide and mental health research, LCA has been used to characterize distinct profiles of risk factors such as suicidal ideation, self-harm behaviors, and psychiatric symptom clusters, highlighting meaningful heterogeneity within at-risk populations and providing clinically interpretable subgroup insights [3, 4]. Prior person-centered studies in self-harm and suicidal behavior populations have similarly identified empirically derived subgroups with differing psychosocial and clinical profiles, demonstrating that heterogeneity in suicide-related phenotypes can be meaningfully captured through such approaches [5].

Moreover, the characteristics and structure of the training data influence both the performance and interpretability of supervised machine learning models [6]. Given the heterogeneous nature of suicide risk, unsupervised exploratory approaches such as LCA may provide complementary insight into underlying population-level patterns of shared risk factors that are not fully captured by individual-level prediction models.

Despite the use of latent class analysis in identifying heterogeneous subgroups related to suicidality, and the use of machine learning approaches to suicide risk assessment [7, 8], to our knowledge, no prior studies have directly examined the correspondence between latent class–derived risk profiles and machine learning–based feature importance, which reflect population-level and individual-level characterizations of suicide risk, respectively. Recent evidence suggests that the clinical utility of machine learning models may be enhanced by complementary approaches that improve interpretability and situate individual-level predictors within broader population-level risk structures [9].

Building on this literature, the present study applies latent class analysis to a publicly available clinical dataset to identify distinct subgroups of individuals with shared patterns of suicide-related characteristics. We then compare these subgroup profiles with findings from a previously published explainable artificial intelligence– based suicide risk assessment model.

We hypothesized that latent class analysis would identify subgroups defined by common patterns of demographic, psychosocial, and clinical features. These latent subgroups were expected to complement machine learning–based suicide risk assessment by providing population-level context that supports and enriches individual-level interpretations, thereby offering a more comprehensive framework for understanding variation in suicide-related outcomes.

## Methods

### Data Sources

This study involved a secondary analysis of a publicly available, anonymized dataset originally collected at Colombo South Teaching Hospital, Kalubowila, Sri Lanka. The dataset was released as part of a software development project conducted in the United Kingdom and is freely accessible for research purposes [10] through a GitHub repository (https://github.com/dinisurunisal/Suicide-Risk-Prediction-Project/blob/master/DataScience/AlgorithmComparison/Test-Data-10.csv). The dataset comprises 1000 participants, with equal representation of individuals who died by suicide and those who did not. Participant ages ranged from 10 to 98 years. Detailed characteristics of the dataset are presented in Table 1.

**Table 1.**
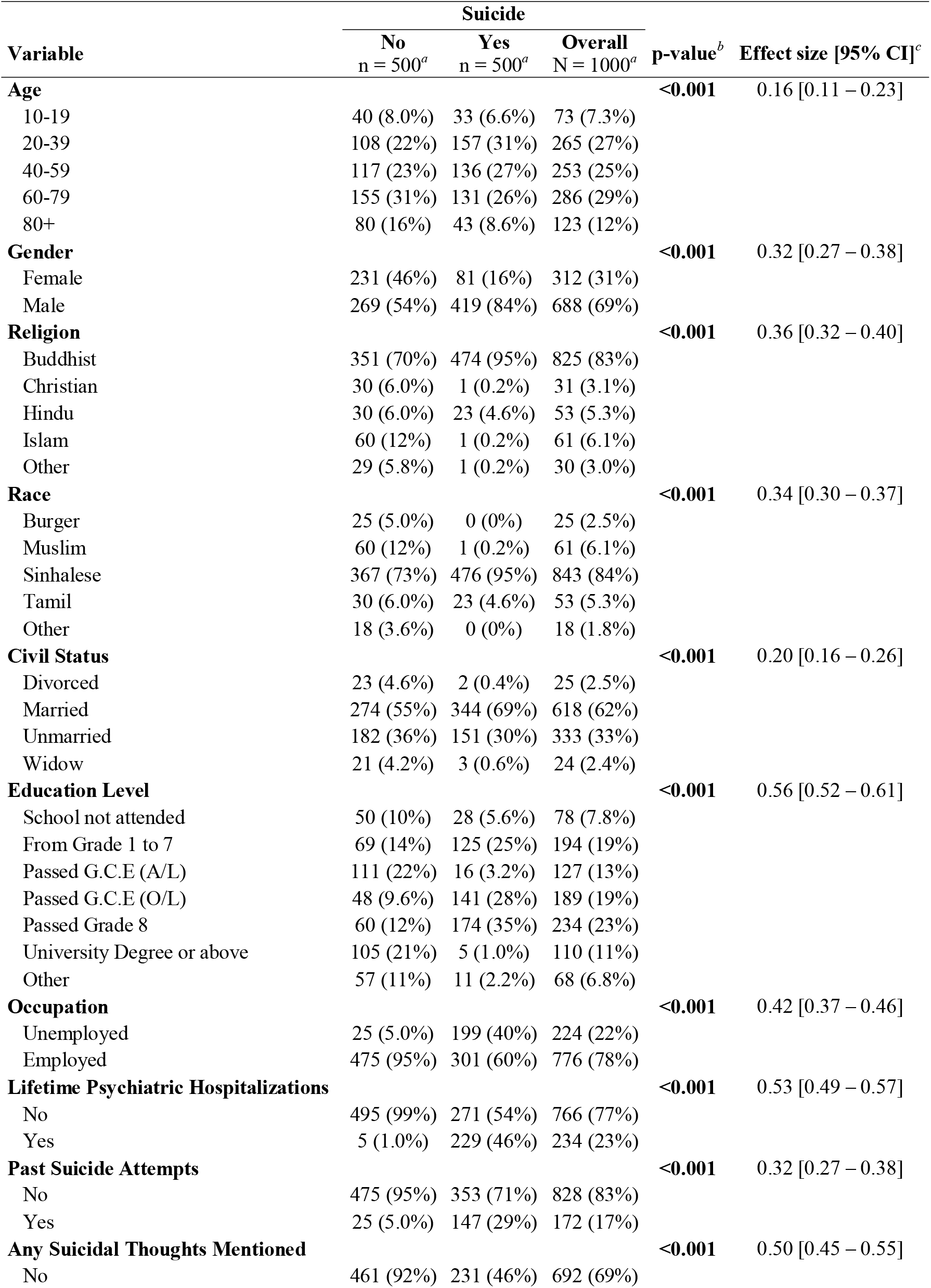
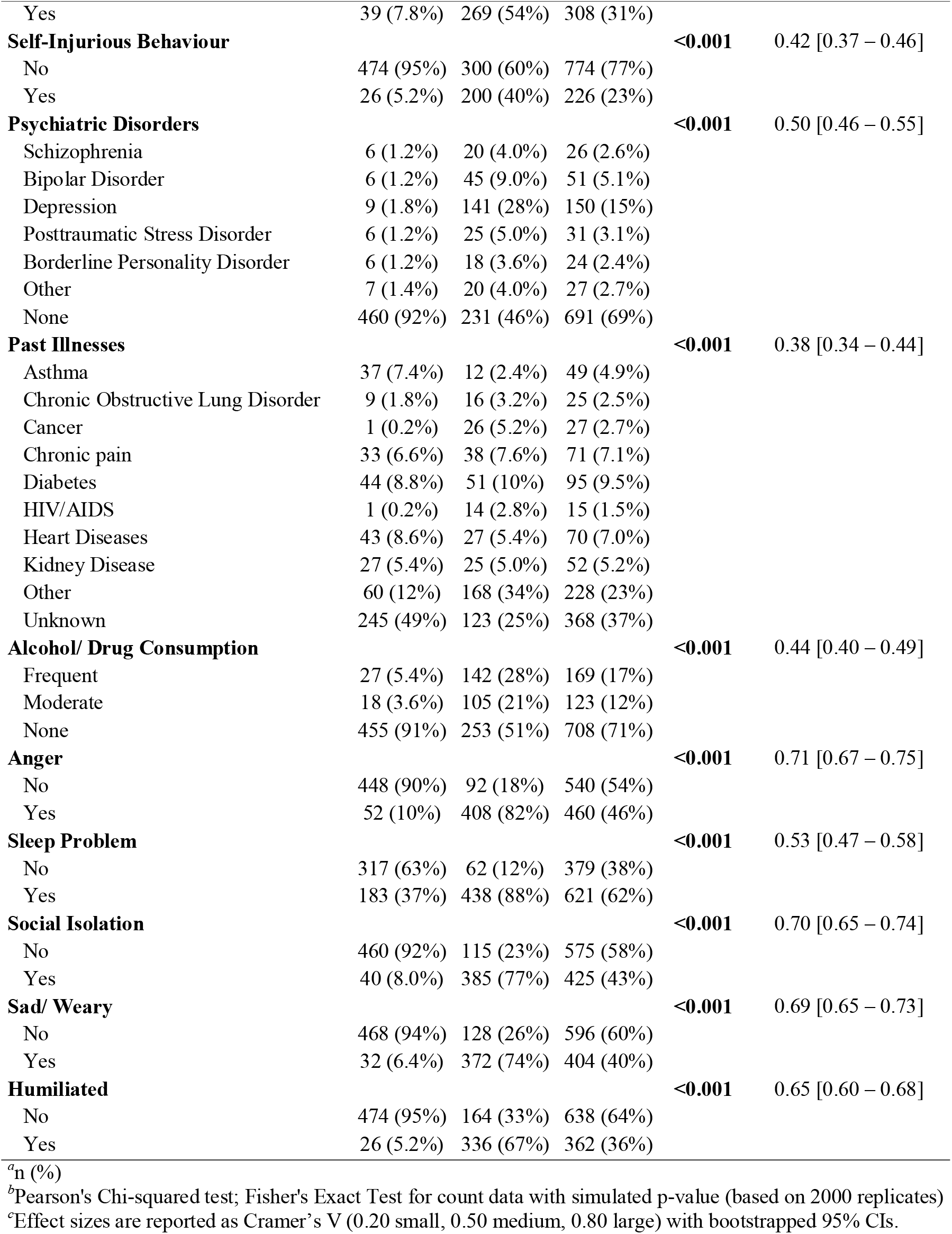
Baseline characteristics of participants by suicide outcome (N = 1000)

The same dataset has previously been analyzed in a supervised machine learning and explainable artificial intelligence–based suicide risk assessment study [10]. In the present study, the dataset was reanalyzed using latent class analysis, and the resulting subgroup patterns were compared descriptively with findings reported in the prior risk assessment study.

The dataset contains sociodemographic, clinical, and psychosocial variables relevant to suicide risk assessment, but no personal identifiable information. As this study involved only secondary analysis of fully de-identified and publicly available data, formal ethical approval and individual informed consent were not required.

### Variables of Interest

The predictors selected in the previous ML–based suicide risk assessment study were used to ensure a robust comparison. Two main modifications were applied for LCA. First, the age variable was categorized into discrete groups rather than treated as continuous, as LCA requires categorical predictors. Second, due to the large number of categories in the occupation variable, it was dichotomized into employed and unemployed to facilitate model estimation. All other variables were used without modification.

The variable indicating suicide outcome (no/yes) was not included in the latent class analysis. Instead, this variable was examined after class assignment to characterize the identified latent subgroups and to assess how the classes differed with respect to the observed outcome.

### Statistical Analysis

Descriptive statistics were used to summarize the demographic, clinical, and psychosocial characteristics of the 1000 participants. Differences between suicide outcome groups (no/yes) were evaluated using p-values and Cramer’s V effect sizes with bootstrapped 95% confidence intervals to quantify the significance and magnitude of observed differences. Categorical variables were compared using the Chi-square test or Fisher’s exact test for sparse categories. A two-sided p-value ≤ 0.05 was considered statistically significant.

Latent class analysis provides a probabilistic, population-level approach by classifying individuals into unobserved subgroups based on shared patterns of risk factors, while accounting for uncertainty in class membership [11, 12]. LCA was used to identify clinically meaningful subgroups reflecting heterogeneity in suicide prevalence. LCA was examined alongside a ML–based suicide risk assessment method, which typically generates non-probabilistic, individual-level risk estimates, to explore similarities and differences in how suicide risk is characterized at the population and individual levels.

Latent class analyses were conducted using R version 4.3.1 with the poLCA package [13]. Models were estimated beginning with a 2-class solution, with the number of classes incrementally increased up to 10 to identify the optimal solution. The upper limit of 10 classes was selected to balance statistical fit with clinical interpretability. To reduce the risk of local maxima, each model was estimated using 100 random starting values.

Model fit was evaluated using the consistent Akaike information criterion (cAIC) and the Bayesian information criterion (BIC), supplemented by the Lo–Mendell–Rubin likelihood ratio test (LMR-LRT), which compares a model with k classes to a model with k − 1 classes [14]. The final model was selected primarily based on the lowest cAIC and BIC values, with additional consideration of class interpretability to ensure clinically meaningful subgroup separation. Classification quality was assessed using entropy, with values greater than 0.80 indicating well-defined class membership [15]. Model evaluation metrics, including class sizes and predicted probability of membership, are reported in Table 2. For each latent class, the prevalence of suicide and corresponding 95% confidence intervals were estimated using Wilson binomial intervals and are presented in Table 3. Variable distributions across classes are provided in Table 4, and the corresponding class profiles for selected variables are shown in Fig 1.

**Fig 1.**
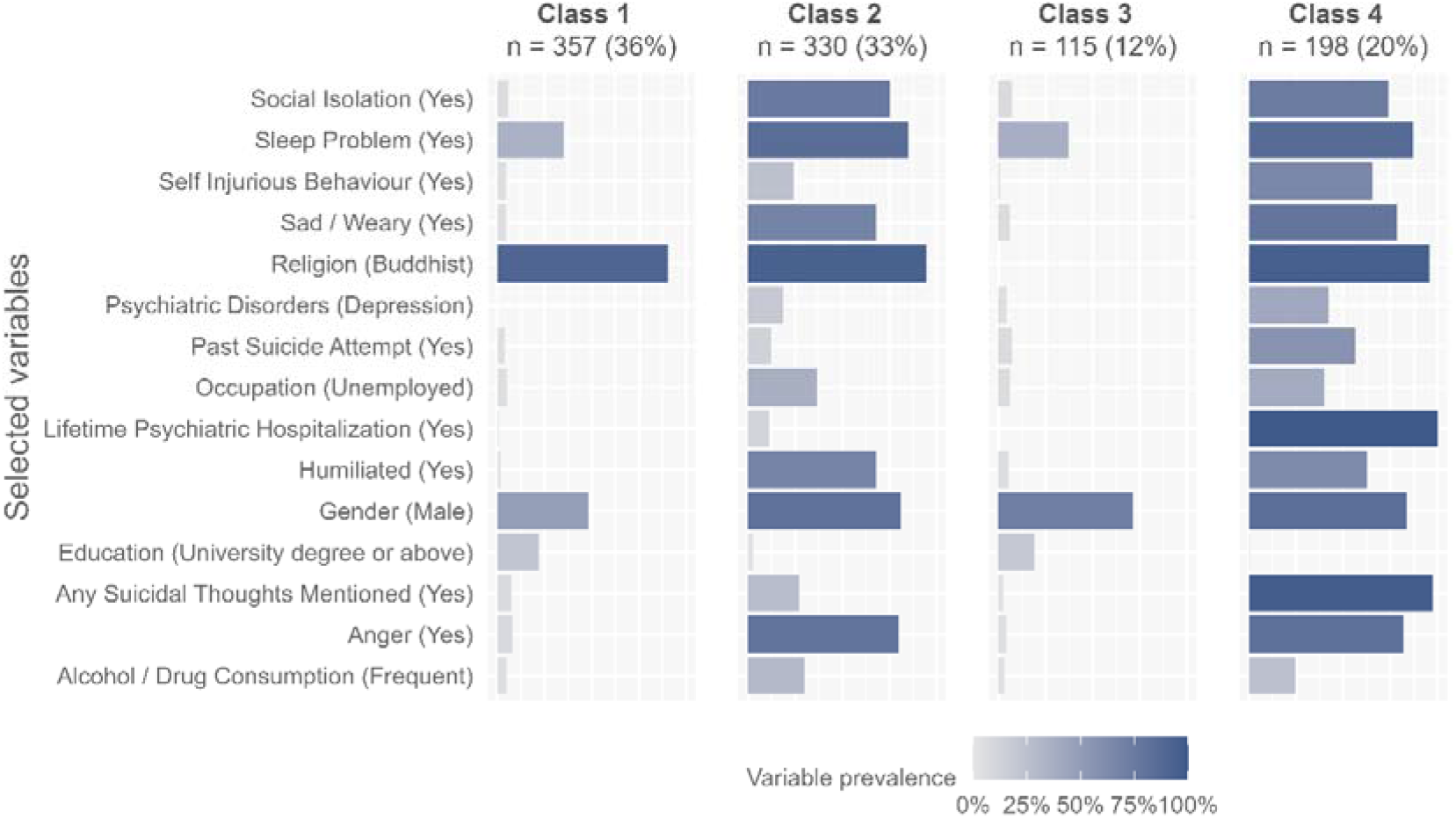
The distributions of variables across classes. Prevalence of suicide in each latent class with 95% confidence intervals: Class 1, 0.6% [0.2–2.0]; Class 2, 91.2% [87.7–93.8]; Class 3, 0.9% [0.2–4.8]; Class 4, 99.0% [96.4–99.7]

## Results

### Descriptive Statistics

The sample included 1000 participants, with equal numbers of individuals dead by suicide (n = 500, 50%) and not dead by suicide. Participants were predominantly male (69%) and Buddhist (83%), with most aged 20–79 years. Significant differences between groups were observed across sociodemographic, clinical, and psychosocial variables (all p < 0.001). Individuals with suicide had higher prevalence of previous psychiatric hospitalization (46% vs 1%), past suicide attempts (29% vs 5%), suicidal thoughts (54% vs 8%), self-injurious behavior (40% vs 5%), and psychiatric disorders such as depression, bipolar disorder, posttraumatic stress disorder (PTSD), and borderline personality disorders (BPD). They also reported higher rates of psychosocial difficulties, including anger, sleep problems, social isolation, and feelings of sadness or humiliation (Cramer’s V range = 0.42–0.71).

### Latent Class Analysis

Latent class analysis identified a four-class solution as the best representation of the data. This solution was supported by fit indices in Table 2, including the lowest BIC (28398) and cAIC (28597), high entropy (0.95), and a significant improvement over the three-class model according to the Lo–Mendell–Rubin likelihood ratio test (p < 0.001). Models with more than four classes yielded smaller class sizes (<115 participants) and lower interpretability. Based on these criteria, the four-class model was retained for further analysis.

Two classes exhibited extremely low suicide prevalence. Class 1 (n = 357, 36%) showed a suicide prevalence of 0.6% (95% CI: 0.2–2.0) and was characterized by low levels of emotional distress, including anger, sadness, social isolation, sleep problems, and humiliation. This class also demonstrated minimal psychiatric morbidity, low substance use, and a high proportion of employed individuals (95%), reflecting a higher level of social, mental and occupational functioning. Class 1 had the highest proportion of female participants (51.6%). Class 3 (n = 115, 12%) similarly demonstrated low suicide prevalence (0.9%, 95% CI: 0.2–4.8) but differed from Class 1 in its clinical profile. Class 3 was characterized by an older age distribution, higher prevalence of somatic conditions such as chronic pain, but low levels of psychosocial distress. Occupation rate was high also in this class (94% employed), but females comprised only 28.7% of participants. The two suicide low-prevalence classes also differed in religion, such as the predominance of Buddhists in Class 1 (90.5%) versus the majority of Muslims (52.3%), Hindus (25.9%), and Christians (21.8%) in Class 3, further supporting their distinction as separate latent phenotypes rather than a single homogeneous group.

In contrast, two classes exhibited exceptionally high suicide prevalence. Class 2 (n = 330, 33%) had a suicide prevalence of 91.2% (95% CI: 87.7–93.8) and was characterized by high levels of emotional and social distress, including anger (80.1%), sadness (68.2%), social isolation (75.6%), sleep problems (84.9%), and humiliation (68.0%). Lifetime psychiatric hospitalization was relatively low in this class (11.3%), and more than half of individuals had no documented psychiatric diagnosis. Employment was lower than in the low-prevalence classes (63.5%), and females accounted for 18.7% of participants. Class 4 (n = 198, 20%) exhibited the highest suicide prevalence at 99.0% (95% CI: 96.4–99.7) and was characterized by universal lifetime psychiatric hospitalization, high prevalence of prior suicide attempts, suicidal thoughts, self-injurious behavior, and psychiatric diagnoses, particularly depression and bipolar disorder. Emotional distress indicators were also highly prevalent, and this class had the lowest employment rate (60.2%) and the smallest female proportion (16.6%). The distribution of the selected variables across classes is shown in Fig 1.

Together, these findings indicate marked heterogeneity in clinical and psychosocial profiles across classes. Occupational status and gender distribution both varied meaningfully across classes, with higher employment and a higher proportion of females observed in low-suicide-prevalence classes, and lower employment and predominantly male composition in high-suicide-prevalence classes. Age, emotional distress, psychiatric disorders, and social factors also contributed to class distinctions, underscoring the multifactorial nature of suicide risk and the value of latent class analysis in identifying clinically and socially relevant subgroups.

### Comparison with Explainable Artificial Intelligence Findings

To contextualize these latent class findings, we compared class-specific profiles with a stratified bar plot (Figure 2) and SHAP-based feature importance patterns (Figure 6-7) reported in the explainable artificial intelligence study “Analysis and evaluation of explainable artificial intelligence on suicide risk assessment” [10]. Figure 2 in Tang et al. [10] shows the distribution of suicidal and non-suicidal records across occupational groups, with unemployed individuals and those in agriculture- and forestry-related jobs exhibiting higher prevalence, consistent with systematic evidence that these occupational groups are at elevated risk of suicide [16]. In our study, high-risk latent classes were predominantly composed of individuals from these same occupational groups (Fig 2).

**Fig 2.**
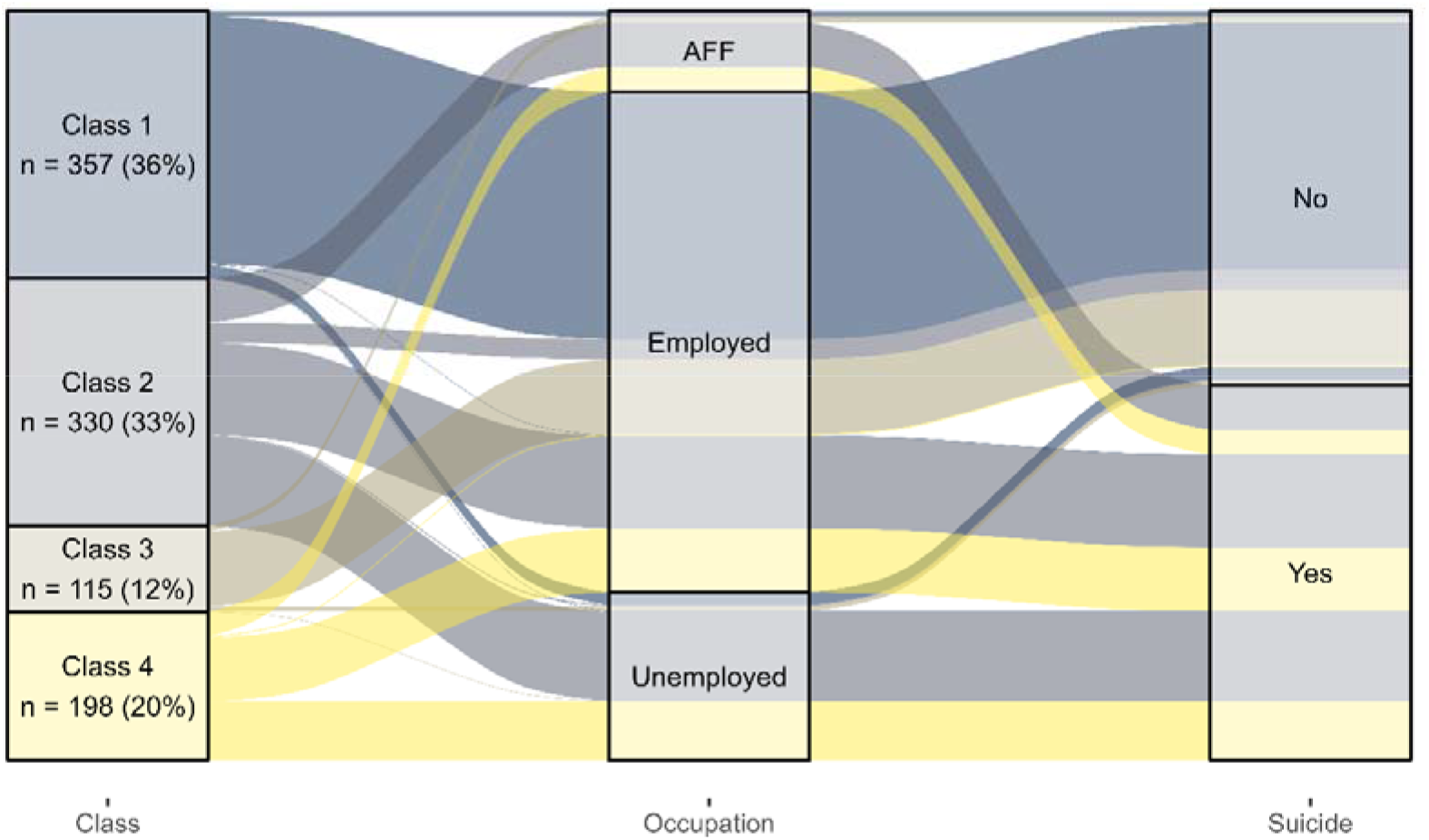
Distribution patterns across latent classes, occupational groups, and suicide outcomes (Occupation was recategorized for visualization as employed, unemployed, and agricultural animal husbandry fisherman & related forestry workers (AFF))

In Tang et al.[10], emotional and psychosocial variables including anger, sadness and social isolation, were identified as the most influential contributors to the model’s predicted probability of suicide, followed by lifetime psychiatric hospitalization, occupation, sleep problems and past suicide attempts (Figure 6). Notably, the emotional and social distress variables ranked highest in the SHAP analysis were consistently elevated across both high-prevalence latent classes identified in the present study. In contrast, lifetime psychiatric hospitalization aligned strongly with only one high-prevalence class (Class 4, 100%), while being largely absent in the other (Class 2, 11.3%). This pattern mirrors the SHAP finding that hospitalization is a strong contributor when present but does not account for all suicide-associated presentations.

The combined results of LCA and SHAP analyses indicate that while lifetime psychiatric hospitalization is a strong discriminative feature when present, high suicide prevalence may also occur in its absence. Emotional and social distress variables were consistently elevated across both high-prevalence classes and ranked highest in SHAP importance, suggesting a shared core set of features associated with suicide across heterogeneous clinical presentations.

Finally, extending the comparison to education, our results closely mirrored those from the ML study. As shown in Figure 7 of Tang et al. [10], lower education levels were associated with higher predicted suicide risk, whereas university education had a protective effect. Similarly, in our latent class analysis, lower-risk classes had a higher proportion of individuals with university education (21.9% and 19.2%), while higher-risk classes had very few (2.6% and 0.6%). Lower educational attainment was more common in higher-risk classes, demonstrating concordance between the approaches.

## Discussion

In this study, LCA revealed substantial heterogeneity in suicide-associated profiles within a cross-sectional sample, identifying two distinct high-prevalence and two low-prevalence classes. By comparing these latent profiles with previously published SHAP-based XAI findings, we demonstrate how unsupervised and supervised approaches provide complementary insights into suicide-associated characteristics.

The SHAP analysis from the prior prediction study highlighted emotional and social distress variables such as anger, sadness and social isolation as the strongest contributors to the model’s predicted probability of suicide [10]. Our latent class findings are highly consistent with this pattern, as these same variables were markedly elevated across both high-prevalence classes, regardless of differences in psychiatric service utilization or diagnostic status. This convergence suggests that emotional and social distress represents a common set of features associated with suicidal behavior across heterogeneous clinical presentations.

At the same time, latent class analysis provides additional nuance not captured by global feature importance alone. While lifetime psychiatric hospitalization was identified as an important contributor in the SHAP analysis, our study shows that this characteristic aligns strongly with only one high-prevalence class (Class 4: 99.0%, 95% CI: 96.4–99.7%). A second high-prevalence class (Class 2: 91.2%, 95% CI: 87.7–93.8%) exhibited similarly elevated suicide prevalence despite minimal prior psychiatric hospitalization and fewer documented diagnoses. This suggests that psychiatric hospitalization characterizes a specific subgroup of individuals with high prevalence of suicidal behaviors, but not all such individuals. Notably, Class 2 included a substantial proportion of older adults (36.6% aged ≥60 years). This pattern aligns with register-based evidence from a different national context showing that a significant proportion of individuals who die by suicide had no prior psychiatric hospitalization and were more likely to be older [17].

The identification of two low-prevalence classes further illustrates the added value of latent class analysis. Although both classes exhibited minimal prevalence of suicidal behaviors, they differed substantially in age distribution, medical burden, psychosocial distress, religious composition and gender distribution. Notably, Class 1 had the highest proportion of females and employed individuals, whereas Class 3, although also low prevalence, differed in age and medical burden. In contrast, both high-prevalence classes were predominantly male with lower employment proportions. Several factors likely contribute to the extremely low suicide prevalence observed in Class 3. Unlike Class 1, none of the individuals in this class were Buddhist, and the majority (75%) belonged to religions such as Islam or Christianity, which may have more restrictive views on suicide. This class also included no Tamil participants, an ethnic group with historically higher suicide rates in Sri Lanka [18], and members had very low rates of prior self-injurious behavior and minimal alcohol or drug consumption. Together, these sociocultural and functional characteristics provide insight into why this subgroup exhibits a relatively protected profile despite the broader population risk factors. These distinctions highlight that low prevalence of suicidal behaviors may arise from multiple phenotypes, and that demographic and sociocultural factors, such as gender and religion, are important characteristics differentiating these latent profiles. Similarly, occupational status may reflect psychosocial and functional stability, with higher employment observed in low-prevalence classes and lower employment in high-prevalence classes. This aligns with prior evidence indicating that unemployment is associated with elevated suicide risk, particularly among males [19].

Together, these findings support the notion that latent class analysis and explainable artificial intelligence serve complementary roles. SHAP-based approaches identify globally influential features associated with suicidal behaviors within predictive models, while latent class analysis delineates distinct subgroups characterized by different constellations of those features, including demographic and functional characteristics such as gender and employment. Integrating insights from both approaches offers a more comprehensive understanding of heterogeneity in suicide-associated characteristics than either method alone.

Given the cross-sectional nature of the data, these findings should be interpreted as identifying patterns and associations rather than temporal or causal pathways. Nevertheless, the convergence between latent class profiles, explainable artificial intelligence findings, and demographic and occupational patterns strengthens confidence in the clinical relevance of the identified emotional, social, and functional characteristics.

### Strengths and Limitations

A key strength of this study is that it complements existing XAI-based suicide risk assessment models by providing a probabilistic, population-level perspective that identifies clinically meaningful subgroups. The fully reproducible methodology and use of a publicly available dataset enhance transparency and enable external validation, while the integration of statistical rigor with clinical insight provides a comprehensive framework for understanding suicide risk beyond individual-level prediction models.

This study has some limitations that should be considered. It is based on a single-site sample from Sri Lanka, which may limit generalizability, and the cross-sectional design precludes assessment of temporal changes in suicide risk. Additionally, the local independence assumption of LCA was not fully met, although prior work suggests this has minimal impact on class assignment [20]. Despite these limitations, the study provides a complementary, probabilistic framework that identifies clinically meaningful subgroups and enhances understanding of suicide risk beyond individual-level prediction models.

## Conclusions

In this study, latent class analysis identified distinct psychosocial and clinical profiles with markedly different suicide prevalence, including two high-prevalence classes characterized by divergent patterns of psychiatric hospitalization.

An important direction for future research is the integration of latent class membership into supervised classification models. Because latent class analysis captures higher-order patterns across psychosocial and clinical variables, class membership may serve as a parsimonious representation of complex phenotypic profiles. Incorporating latent class labels into explainable artificial intelligence frameworks may enhance interpretability and potentially improve classification performance by linking individual predictions to broader clinical phenotypes.

## Data Availability

All data produced in the present work are contained in the manuscript

https://github.com/dinisurunisal/Suicide-Risk-Prediction-Project/blob/master/DataScience/AlgorithmComparison/Test-Data-10.csv

https://github.com/busenurk/suicide-lca-xai-comparison

## Availability of data and material

The data used in this study are publicly available in an open-access repository at: https://github.com/dinisurunisal/Suicide-Risk-Prediction-Project/blob/master/DataScience/AlgorithmComparison/Test-Data-10.csv

## Code availability

The R code used in this study is publicly available at Busenur Kizilaslan’s GitHub repository (https://github.com/busenurk/suicide-lca-xai-comparison)

## Compliance with ethical standards

## Appendix A

**Table 2.** Fit indices and classification quality measures for latent class models.

| Class | BIC<sup>a</sup> | cAIC<sup>b</sup> | Entropy | Smallest Class Count | LMR_p<sup>c</sup> | Predicted Probability of Membership |  |  |  |  |  |  |  |  |  |
| --- | --- | --- | --- | --- | --- | --- | --- | --- | --- | --- | --- | --- | --- | --- | --- |
|  |  |  |  |  |  | 1 | 2 | 3 | 4 | 5 | 6 | 7 | 8 | 9 | 10 |
| 2 | 28749 | 28848 | 0.96 | 470 | - | 0.47 | 0.53 |  |  |  |  |  |  |  |  |
| 3 | 28508 | 28657 | 0.97 | 115 | <0.001 | 0.52 | 0.12 | 0.36 |  |  |  |  |  |  |  |
| 4 | **28398** | **28597** | **0.95** | **115** | **<0.001** | **0.36** | **0.34** | **0.11** | **0.19** |  |  |  |  |  |  |
| 5 | 28489 | 28738 | 0.93 | 93 | <0.001 | 0.28 | 0.11 | 0.16 | 0.35 | 0.09 |  |  |  |  |  |
| 6 | 28603 | 28902 | 0.94 | 28 | <0.001 | 0.35 | 0.27 | 0.15 | 0.09 | 0.11 | 0.03 |  |  |  |  |
| 7 | 28741 | 29090 | 0.94 | 24 | <0.001 | 0.15 | 0.05 | 0.09 | 0.06 | 0.27 | 0.02 | 0.35 |  |  |  |
| 8 | 28915 | 29314 | 0.92 | 24 | <0.001 | 0.30 | 0.06 | 0.27 | 0.13 | 0.06 | 0.1 | 0.05 | 0.02 |  |  |
| 9 | 29115 | 29564 | 0.95 | 24 | <0.001 | 0.13 | 0.20 | 0.35 | 0.02 | 0.05 | 0.05 | 0.06 | 0.06 | 0.06 |  |
| 10 | 29296 | 29795 | 0.91 | 25 | <0.001 | 0.12 | 0.06 | 0.07 | 0.11 | 0.06 | 0.08 | 0.02 | 0.29 | 0.13 | 0.05 |

<sup>a</sup>BIC: Bayesian Information Criterion
<sup>b</sup>cAIC: Consistent Akaike Information Criterion
<sup>c</sup>LMR\_p: Lo-Mendell-Rubin likelihood ratio test p value
The bolded class was identified as the optimal model, considering both statistical fit criteria and interpretability.

**Table 3.** Class-specific prevalence of suicide.

| Class | Suicide (n) | Total (n) | Prevalence % [95% CI] |
| --- | --- | --- | --- |
| 1 | 2 | 357 | 0.6 [0.2-2] |
| 2 | 301 | 330 | 91.2 [87.7-93.8] |
| 3 | 1 | 115 | 0.9 [0.2-4.8] |
| 4 | 196 | 198 | 99 [96.4-99.7] |

**Table 4.** Comparison of observed and predicted variable prevalence across latent classes.

| **Variable** | **Overall**<br>**Observed**<br>**Prevalence**<br>**(N = 1000)** | **Class 1**<br>**(n = 357,**<br>**36%)** | **Class 2**<br>**(n = 330,**<br>**33%)** | **Class 3**<br>**(n = 115,**<br>**12%)** | **Class 4**<br>**(n = 198,**<br>**20%)** |
| --- | --- | --- | --- | --- | --- |
| **Age** |  |  |  |  |  |
| 10-19 | 7.3% | 7.8% | 5.5% | 5.2% | 10.7% |
| 20-39 | 26.5% | 22.2% | 32.6% | 19.2% | 28.2% |
| 40-59 | 25.3% | 21.7% | 25.3% | 31.4% | 28.4% |
| 60-79 | 28.6% | 32.4% | 27% | 26.7% | 25.3% |
| 80+ | 12.3% | 15.9% | 9.6% | 17.4% | 7.3% |
| **Gender** |  |  |  |  |  |
| Female | 31.2% | 51.6% | 18.7% | 28.7% | 16.6% |
| Male | 68.8% | 48.4% | 81.3% | 71.3% | 83.4% |
| **Religion** |  |  |  |  |  |
| Buddhist | 82.5% | 90.5% | 94.6% | 0% | 95.8% |
| Christian | 3.1% | 1.4% | 0% | 21.8% | 0.5% |
| Hindu | 5.3% | 0% | 5.1% | 25.9% | 3.2% |
| Islam | 6.1% | 0% | 0.3% | 52.3% | 0% |
| Other | 3% | 8.1% | 0% | 0% | 0.5% |
| **Race** |  |  |  |  |  |
| Burger | 2.5% | 0% | 0% | 21.8% | 0% |
| Muslim | 6.1% | 0% | 0.3% | 52.3% | 0% |
| Sinhalese | 1.8% | 5% | 0% | 0% | 0% |
| Tamil | 84.3% | 95% | 94.6% | 0% | 96.8% |
| Other | 5.3% | 0% | 5.1% | 25.9% | 3.2% |
| **Civil Status** |  |  |  |  |  |
| Divorced | 2.5% | 6.4% | 0.4% | 0% | 0.5% |
| Married | 61.8% | 55.2% | 69.9% | 54.8% | 64.2% |
| Unmarried | 33.3% | 34% | 28.6% | 42.6% | 34.8% |
| Widow | 2.4% | 4.5% | 1.2% | 2.6% | 0.5% |
| **Education Level** |  |  |  |  |  |
| School not attended | 7.8% | 10.5% | 4% | 10.6% | 7.9% |
| From Grade 1 to 7 | 19.4% | 12.4% | 24.9% | 14% | 26% |
| Passed G.C.E (A/L) | 12.7% | 22.5% | 3.9% | 23.5% | 3.5% |
| Passed G.C.E (O/L) | 18.9% | 9.3% | 25.6% | 11.3% | 29.4% |
| Passed Grade 8 | 23.4% | 11.2% | 36.7% | 10.9% | 30.3% |
| University Degree or above | 11% | 21.9% | 2.6% | 19.2% | 0.6% |
| Other | 6.8% | 12.3% | 2.3% | 10.5% | 2.3% |
| **Occupation** |  |  |  |  |  |
| Unemployed | 22.4% | 5% | 36.5% | 6% | 39.8% |
| Employed | 77.6% | 95% | 63.5% | 94% | 60.2% |
| **Lifetime Psychiatric Hospitalizations** |  |  |  |  |  |
| No | 76.6% | 99.1% | 88.7% | 100% | 0% |
| Yes | 23.4% | 0.9% | 11.3% | 0% | 100% |
| **Past Suicide Attempts** |  |  |  |  |  |
| No | 82.8% | 96.1% | 87.6% | 93.1% | 43.6% |
| Yes | 17.2% | 3.9% | 12.4% | 6.9% | 56.4% |
| **Any Suicidal Thoughts Mentioned** |  |  |  |  |  |
| No | 69.2% | 92.7% | 72.9% | 97.4% | 2.5% |

| Variable | Overall<br>Observed<br>Prevalence<br>(N = 1000) | Class 1<br>(n = 357,<br>36%) | Class 2<br>(n = 330,<br>33%) | Class 3<br>(n = 115,<br>12%) | Class 4<br>(n = 198,<br>20%) |
| --- | --- | --- | --- | --- | --- |
| Yes | 30.8% | 7.3% | 27.1% | 2.6% | 97.5% |
| **Self-Injurious Behaviour** |  |  |  |  |  |
| No | 77.4% | 95.4% | 75.5% | 99.1% | 34.5% |
| Yes | 22.6% | 4.6% | 24.5% | 0.9% | 65.5% |
| **Psychiatric Disorders** |  |  |  |  |  |
| Schizophrenia | 2.6% | 0.3% | 3.8% | 1.7% | 5.2% |
| Bipolar Disorder | 5.1% | 0.6% | 8.2% | 0.9% | 10.4% |
| Depression | 15% | 0.3% | 18.7% | 4.3% | 42.1% |
| Posttraumatic Stress Disorder | 3.1% | 0.6% | 4.3% | 0% | 7.6% |
| Borderline Personality Disorder | 2.4% | 1.2% | 3.2% | 0% | 4.6% |
| Other | 2.7% | 1.3% | 4.8% | 0.9% | 2.7% |
| None | 69.1% | 95.7% | 57% | 92.2% | 27.3% |
| **Past Illnesses** |  |  |  |  |  |
| Asthma | 4.9% | 7.9% | 2.7% | 6.9% | 1.9% |
| Chronic Obstructive Lung<br>Disorder | 2.5% | 2% | 3.3% | 1.7% | 2.5% |
| Cancer | 2.7% | 0% | 3.7% | 0.9% | 7% |
| Chronic pain | 7.1% | 3.9% | 7.6% | 14.7% | 7.7% |
| Diabetes | 9.5% | 7.3% | 11.3% | 9.6% | 10.4% |
| HIV/AIDS | 1.5% | 0% | 2.7% | 0% | 3% |
| Heart Diseases | 7% | 10% | 4.2% | 6.1% | 6.8% |
| Kidney Disease | 5.2% | 6.4% | 4.8% | 3.5% | 4.7% |
| Other | 22.8% | 12.4% | 36.9% | 7.9% | 26.4% |
| Unknown | 36.8% | 50% | 22.8% | 48.7% | 29.7% |
| **Alcohol/ Drug Consumption** |  |  |  |  |  |
| Frequent | 16.9% | 4.9% | 29.8% | 3.2% | 24.7% |
| Moderate | 12.3% | 3.4% | 19.3% | 2.8% | 22.2% |
| None | 70.8% | 91.6% | 50.9% | 94% | 53.1% |
| **Anger** |  |  |  |  |  |
| No | 54% | 92.1% | 19.9% | 95.9% | 17.9% |
| Yes | 46% | 7.9% | 80.1% | 4.1% | 82.1% |
| **Sleep Problem** |  |  |  |  |  |
| No | 37.9% | 64.8% | 15.1% | 62.7% | 13% |
| Yes | 62.1% | 35.2% | 84.9% | 37.3% | 87% |
| **Social Isolation** |  |  |  |  |  |
| No | 57.5% | 94.3% | 24.4% | 92.9% | 26% |
| Yes | 42.5% | 5.7% | 75.6% | 7.1% | 74% |
| **Sad/ Weary** |  |  |  |  |  |
| No | 59.6% | 95.4% | 31.8% | 93.9% | 21.3% |
| Yes | 40.4% | 4.6% | 68.2% | 6.1% | 78.7% |
| **Humiliated** |  |  |  |  |  |
| No | 63.8% | 98.1% | 32% | 94.8% | 37.1% |
| Yes | 36.2% | 1.9% | 68% | 5.2% | 62.9% |

